# Fatal breakthrough infection after anti-BCMA CAR-T therapy highlights suboptimal immune response to SARS-CoV-2 vaccination in myeloma patients

**DOI:** 10.1101/2021.05.15.21256814

**Authors:** Adolfo Aleman, Oliver Van Oekelen, Bhaskar Upadhyaya, Sarita Agte, Katerina Kappes, Katherine Beach, Komal Srivastava, Charles R. Gleason, PVI study group, Bo Wang, Tarek H. Mouhieddine, Kevin Tuballes, Daniel Geanon, Zenab Khan, Ana S. Gonzalez-Reiche, Harm van Bakel, Nicole W. Simons, Konstantinos Mouskas, Alexander W. Charney, Adeeb Rahman, Seunghee Kim-Schulze, Emilia M. Sordillo, Florian Krammer, Carlos Cordon-Cardo, Nina Bhardwaj, Sacha Gnjatic, Miriam Merad, Brian D. Brown, Larysa Sanchez, Ajai Chari, Sundar Jagannath, Viviana Simon, Ania Wajnberg, Samir Parekh

## Abstract

Severe acute respiratory syndrome coronavirus 2 (SARS-CoV-2) vaccines are highly effective in healthy individuals. Patients with multiple myeloma (MM) are immunocompromised due to defects in humoral and cellular immunity as well as immunosuppressive therapies. The efficacy after two doses of SARS-CoV-2 mRNA vaccination in MM patients is currently unknown. Here, we report the case of a MM patient who developed a fatal SARS-CoV-2 infection after full vaccination while in remission after B cell maturation antigen (BCMA)-targeted chimeric antigen receptor (CAR)-T treatment. We show that the patient failed to generate antibodies or SARS-CoV-2-specific B and T cell responses, highlighting the continued risk of severe coronavirus disease 2019 (COVID-19) in vaccine non-responders. In the largest cohort of vaccinated MM patients to date, we demonstrate that 15.9% lack SARS-CoV-2 spike antibody response more than 10 days after the second mRNA vaccine dose. The patients actively receiving MM treatment, especially on regimens containing anti-CD38 and anti-BCMA, have lower antibody responses compared to healthy controls. Thus, it is of critical importance to monitor this patient population for serological responses. Non-responders may benefit from ongoing public health measures and from urgent study of prophylactic treatments to prevent SARS-CoV-2 infection.

## INTRODUCTION

Severe acute respiratory syndrome coronavirus 2 (SARS-CoV-2) mRNA vaccines are highly efficacious in preventing coronavirus disease 2019 (COVID-19) morbidity and mortality^1,2^. Preliminary reports suggest that the antibody response in MM after the initial dose of SARS-CoV-2 mRNA vaccine is attenuated and delayed compared to healthy controls, but information on response after completion of the full two-dose mRNA vaccine regimen in MM patients is currently lacking^3-5^. Moreover, studies have not yet examined the T cell response to vaccination in MM patients. The kinetics of the vaccine response in MM patients with prior COVID-19 and impact of treatments on vaccine response are also unknown.

Here, we present the clinical course of a MM patient who received both vaccine doses after B cell maturation antigen (BCMA)-targeted chimeric antigen receptor (CAR)-T therapy. Despite vaccination, the patient developed a fatal infection with SARS-CoV-2. We show that this patient did not generate circulating anti-spike IgG antibodies, but notably, that the patient also failed to mount detectable SARS-CoV-2-specific B and T cell responses. We contextualize this case by summarizing serological results from a large cohort of 208 vaccinated MM patients compared with 38 age-matched fully vaccinated healthy controls. Importantly, we report on treatment- and disease-related characteristics associated with the absence of serological responses after two doses of mRNA vaccination (Moderna/Pfizer). Taken together, these observations support monitoring of quantitative SARS-CoV-2 spike antibody levels as well as further study of B and T cell responses in the vulnerable MM population to identify patients that remain at risk for severe COVID-19 despite vaccination.

## RESULTS

Patient A is an IgG kappa MM patient and enrolled in a clinical trial of BCMA-targeted CAR-T cell therapy with a 4-year history of relapsed/refractory myeloma after 5 lines of treatment in the fall of 2020 (Figure 1A). Following apheresis, the patient received lymphodepleting chemotherapy (fludarabine + cyclophosphamide) before CAR-T infusion. Patient A received Pfizer-BioNTech mRNA SARS-CoV-2 vaccination more than three months after CAR-T infusion in accordance with the international guidelines^6^. Blood cell counts (including absolute lymphocyte count) were within normal limits (Supplementary Figure S1), the bone marrow was negative for clonal plasma cells and serologic MM markers were consistent with very good partial response (VGPR) at the time of both vaccine doses. The patient was admitted to the hospital two weeks after receiving the second vaccine dose with progressive dyspnea, hypoxia, and fever. SARS-CoV-2 molecular testing was positive and complete viral genome sequencing revealed the presence of the B.1.1.7 variant. The patient’s oxygen requirements escalated and ultimately advanced to mechanical ventilation. Despite best supportive care, including remdesivir, corticosteroids, high-titer convalescent plasma, broad-spectrum antibacterial and antifungal agents, intravenous immunoglobulin and multiple vasopressors, the patient’s clinical condition deteriorated, and the patient ultimately passed away.

**Figure 1:**
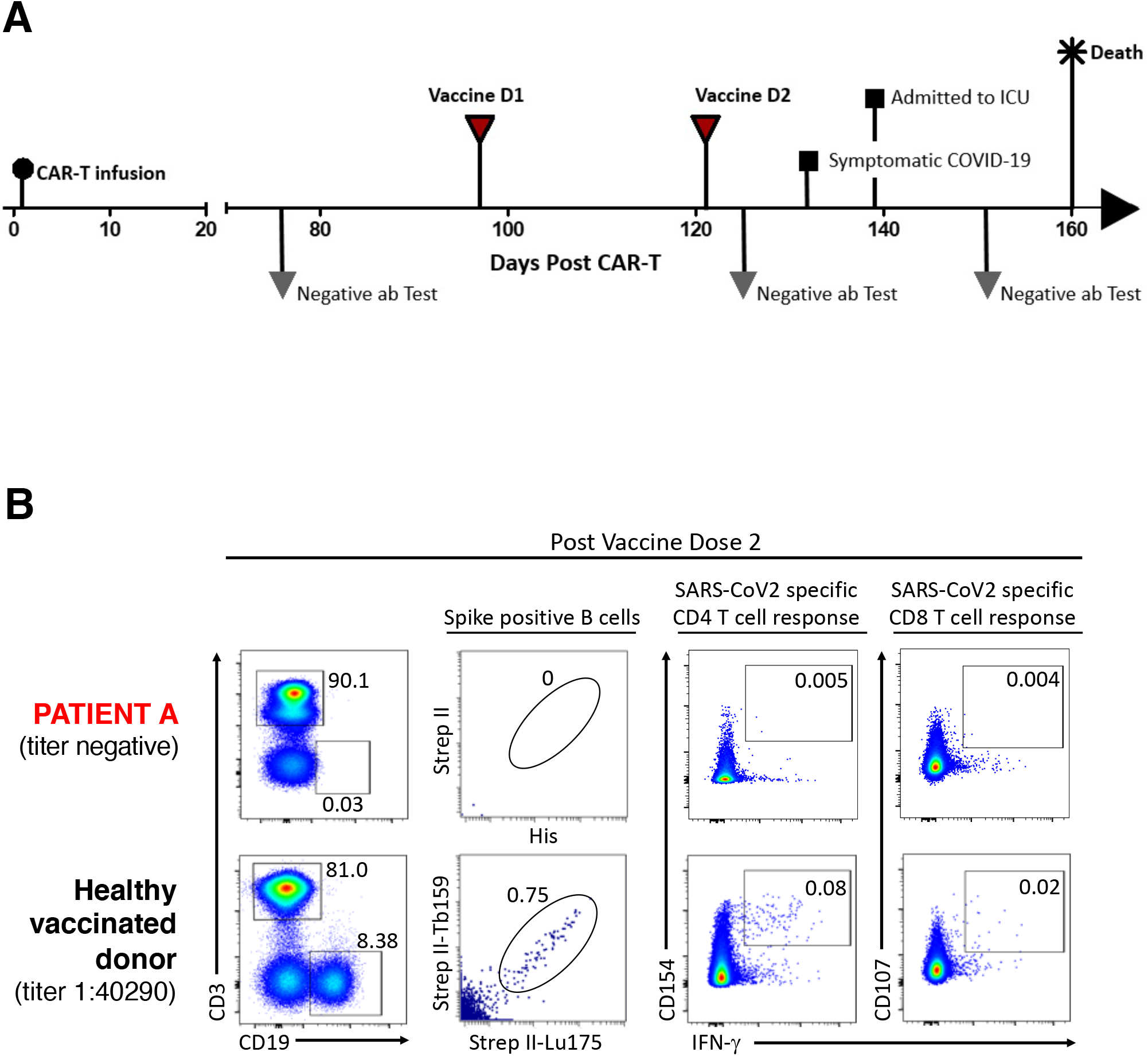
Clinical summary of fatal SARS-CoV-2 infection and absence of spike-specific B cells and SARS-CoV-2-specific T cell responses in a B cell maturation antigen (BCMA) chimeric antigen receptor (CAR)-T patient after two doses of SARS-CoV-2 mRNA vaccine. (A) Clinical event timeline, showing CAR-T infusion (day 0), repeated negative (< 5 AU/mL) SARS-CoV-2 IgG antibody tests (day 76, day 125 and day 151), administration of both doses of the Pfizer-BioNTech mRNA vaccine (day 97 and day 125), development of symptomatic COVID-19 (day 132), admission to the intensive care unit (ICU) (day 139) and death (day 160). (B) Lack of B cell and T cell responses to SARS-CoV-2 peptide stimulations, shown by flow cytometry on peripheral blood mononuclear cells (PBMC) in patient A (top, sample collected post vaccine dose 2) and a healthy vaccinated donor (bottom, sample collected post vaccine dose 2). The leftmost column shows a complete depletion of CD19+ B cells in patient A (% of CD45+ cells shown) in comparison to the healthy donor. Second column shows the absence of SARS-CoV-2 spike-positive (i.e. spike-specific) B cells in Patient compared to a healthy vaccinated donor (% of CD19+ cells shown). The third column shows absence of spike-specific activated CD4+ T cells (CD4+CD154+IFN-γ+) in patient A in comparison to the healthy vaccinated donor (% of CD3+CD4+ T cells shown). The right plots illustrate relative absence of spike-specific activated CD8+ T cells (CD8+CD107+IFN-γ+) in patient A in comparison to the healthy vaccinated donor (% of CD3+CD8+ T cells shown). T cells were stimulated with CD4+ and CD8+ SARS-CoV-2 peptide pools compared to a healthy vaccinated donor with a seropositive antibody titer at the time of sample collection.

We observed the absence of SARS-CoV-2 IgG antibodies following the second vaccine dose in patient A (Figure 1A). Because it has been suggested that T cell responses may offer some protection, even in the absence of circulating antibodies^7,8^, we examined SARS-CoV-2-specific B and T cell responses after vaccination in patient A compared to a healthy donor (Figure 1B). Flow cytometry revealed complete B cell depletion in the peripheral blood of patient A starting immediately following BCMA-targeted therapy and persisting throughout the two-dose vaccination regimen (Figure 1B, Supplementary Figure S1F). We also confirmed the lack of SARS-CoV-2 spike-specific B cells in patient A in contrast to two vaccinated healthy controls and a previously SARS-CoV-2 infected, recovered and non-vaccinated (i.e. COVID-19 convalescent) peripheral blood mononuclear cell (PBMC) donor, using a mass cytometry (CyTOF) assay (Figure 1B, Supplementary Figure S2)

Intracellular cytokine measurement after stimulation with either peptides derived from total viral protein or only from spike protein showed complete absence of CD4 and CD8 T cell response in patient A despite receiving both vaccine doses. In contrast, we observed SARS-CoV-2 peptide-specific T cell responses in a vaccinated healthy control (Figure 2B) as well as in the COVID-19 convalescent PBMC donor (Supplementary Figure S3A-D). Of note, PBMC of patient A yielded robust recall responses to CEFT peptide pool (CMV, EBV, influenza and tetanus toxin) and SEB (Staphylococcal Enteroroxin B superantigen) highlighting the fact that antigen presentation and T cell activation of patient A were not, per se, compromised (Supplementary Figure S3E). T cell responses were independently confirmed using enzyme-linked immunospot (ELISpot) assays (Supplementary Figure S4) where responses to full-length spike protein was tested in addition to SARS-CoV-2 peptide pools. No IFN-γ spot forming cells (SFC) were observed upon stimulation of PBMC from Patient A with SARS-CoV-2 peptides nor full-length spike protein, whereas IFN-γ SFC were readily detected in the PBMC from the vaccinated healthy controls and from the asymptomatic COVID-19 convalescent PBMC donor.

**Figure 2:**
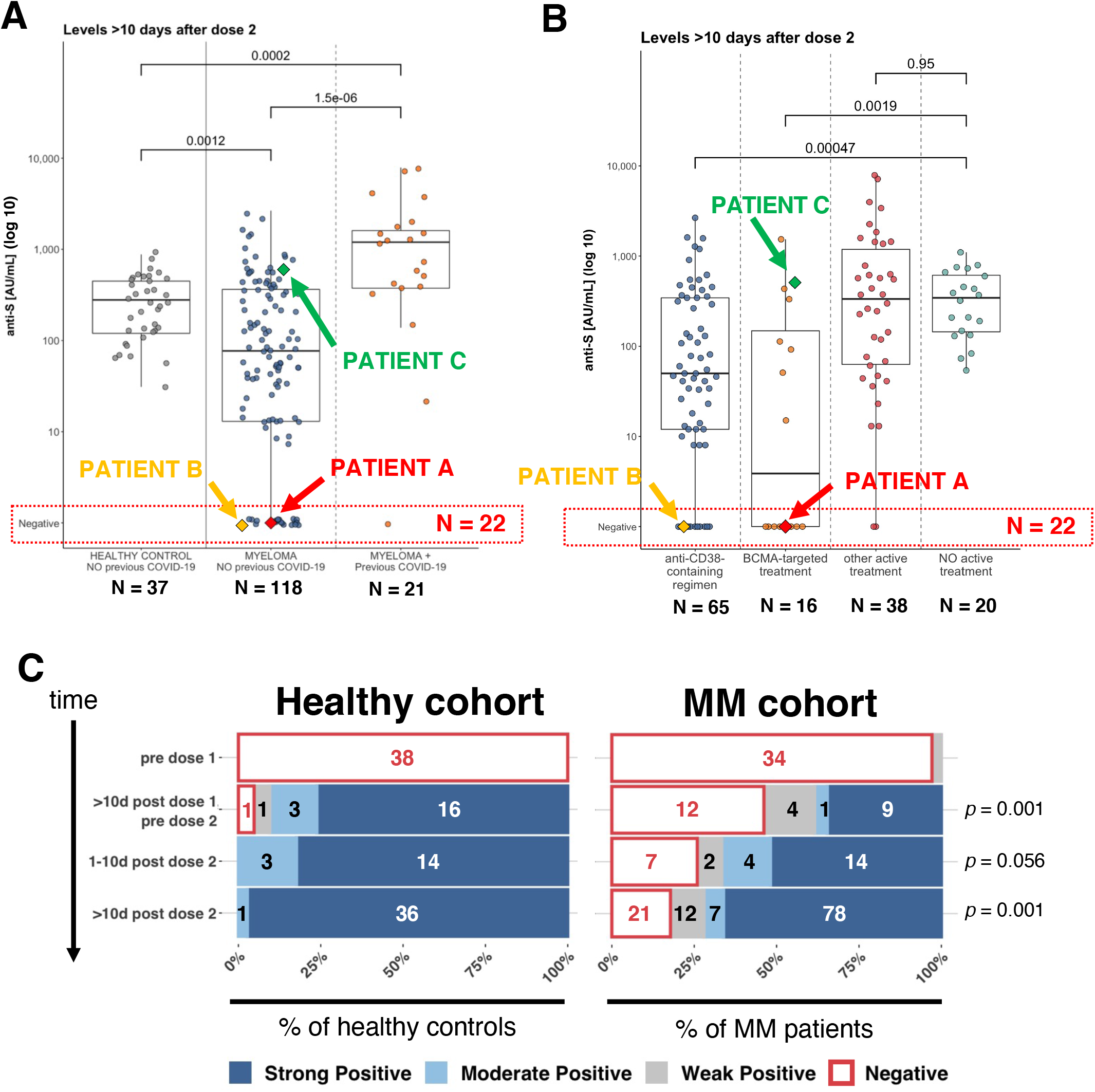
Anti-spike (S) IgG antibody responses after two doses of SARS-CoV-2 mRNA vaccine is delayed and suboptimal or absent in multiple myeloma (MM) patients compared to healthy donors. (A) SARS-CoV-2 anti-S IgG antibody level (shown on log-10 scale) at least 10 days after receiving two doses of SARS-CoV-2 mRNA vaccine in healthy controls without prior COVID-19 infection (gray, left), multiple myeloma (MM) patients without prior COVID-19 infection (blue, center) and MM patients with prior COVID-19 infection (red, right). P-values shown according to non-parametric Mann-Whitney U test. (B) SARS-CoV-2 anti-S IgG antibody level (shown on log-10 scale) at least 10 days after receiving two doses of SARS-CoV-2 mRNA vaccine in MM patients split according to major treatment groups. P-values shown according to the non-parametric Mann-Whitney U test. (C) Qualitative SARS-CoV-2 anti-S IgG antibody measurements of a healthy cohort (left) and a cohort of MM patients (right) showing delayed seroconversion and complete absence of SARS-CoV-2 anti-S IgG antibodies in a subgroup of MM patients. P-values represent comparison between the healthy cohort and MM cohort of the qualitative antibody measurement distribution at the annotated time points using Fisher’s Exact Test on a 2×4 contingency table.

Additional controls for our analysis include a fully vaccinated SARS-CoV-2 seronegative MM patient (patient B) on an anti-CD38 monoclonal antibody-containing regimen who also showed negative IgG titers, lack of SARS-CoV-2-specific B and T cell responses and depleted B cells in the peripheral blood (Supplementary Figures S2-4). In contrast, another MM patient (patient C), who received two doses of mRNA vaccine 18 months after BCMA-targeted CAR-T mounted robust antibody titers and demonstrated B and T cell responses to SARS-CoV-2 peptides (Supplementary Figures S2-4). Notably, patient C recovered B cell numbers prior to vaccination (Supplementary Figure S2). Taken together, these results not only suggest that the absence of B cells is responsible for the failure of the SARS-CoV-2 vaccines to induce a B cell response in the form of SARS-CoV-2 antibodies but also point to the possibility that the lack of B cells is involved in the failure to develop proper T cell responses.

To characterize our finding of persistently negative antibody titers after both SARS-CoV-2 mRNA vaccine doses, we studied serological immune responses in a larger cohort of 208 MM patients enrolled in several institutional review board (IRB)-approved observational studies (see Methods for details). All participants had been diagnosed with myeloma and had received, at least, a first dose of mRNA vaccine (70.2% Pfizer-BioNTech, 24.5% NIH-Moderna). At the time of the analysis, 139/208 MM patients (66.8%) had received both mRNA vaccine doses and had SARS-CoV-2 IgG antibody levels measured at least 10 days after receiving the second dose. Of note, 21/139 fully vaccinated MM patients (15.1%) had a previously documented SARS-CoV-2 infection and were seropositive for SARS-CoV-2 prior to immunization. The median age of the MM cohort is 68 years (range 38-93 years) with 58.7% of participants being male. Additional clinical and disease characteristics are outlined in Table 1.

**TABLE 1.**
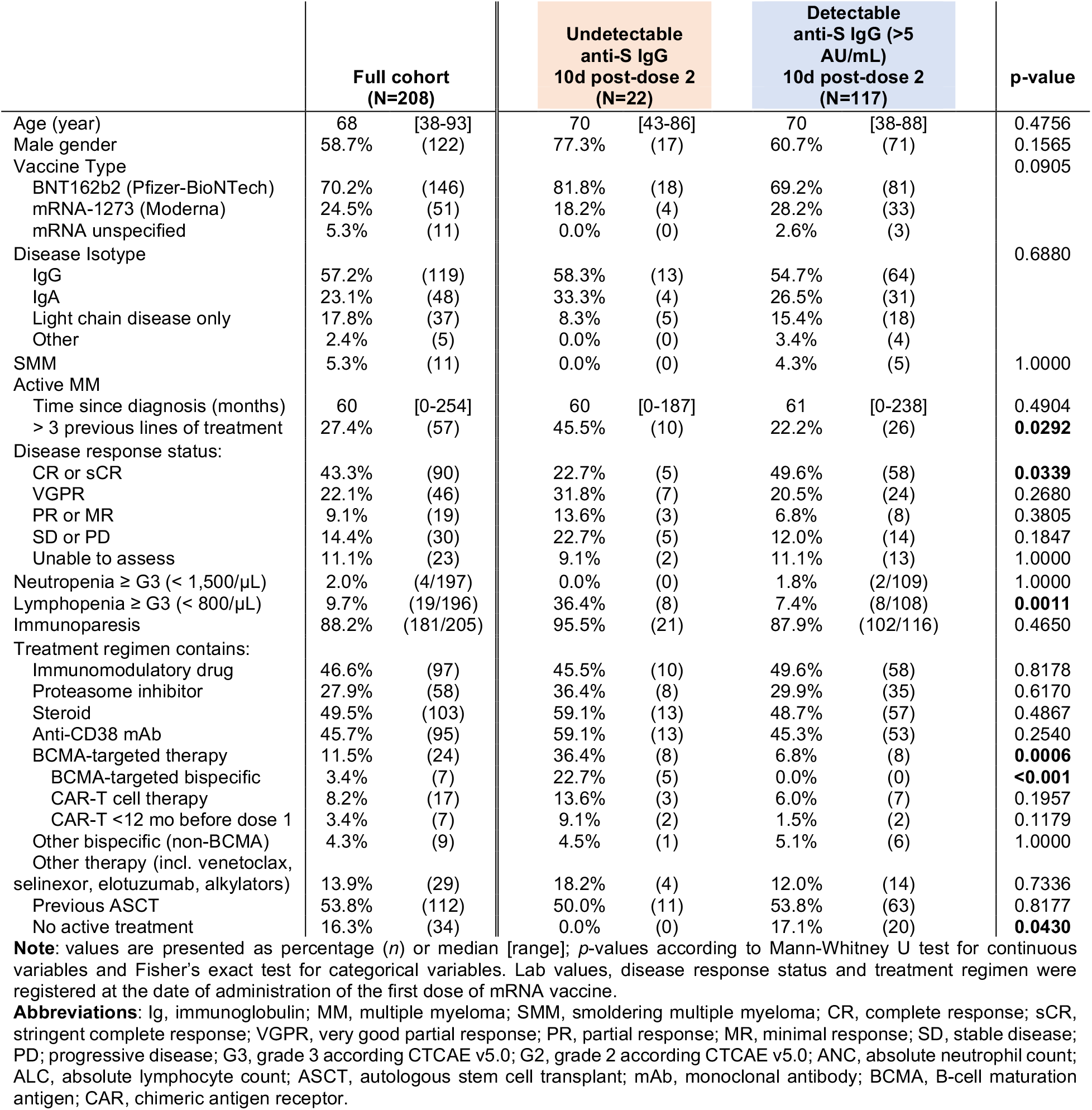
Clinical characteristics of the myeloma cohort and univariate analysis of disease and treatment characteristics associated with absence of antibody levels > 10 days after receiving two doses of SARS-CoV-2 mRNA vaccine.

|  | Full cohort<br>(N=208) |  | Undetectable<br>anti-S IgG<br>10d post-dose 2<br>(N=22) |  | Detectable<br>anti-S IgG (>5<br>AU/mL)<br>10d post-dose 2<br>(N=117) |  | p-value |
| --- | --- | --- | --- | --- | --- | --- | --- |
| Age (year) | 68 | [38-93] | 70 | [43-86] | 70 | [38-88] | 0.4756 |
| Male gender | 58.7% | (122) | 77.3% | (17) | 60.7% | (71) | 0.1565 |
| Vaccine Type |  |  |  |  |  |  | 0.0905 |
| BNT162b2 (Pfizer-BioNTech) | 70.2% | (146) | 81.8% | (18) | 69.2% | (81) |  |
| mRNA-1273 (Moderna) | 24.5% | (51) | 18.2% | (4) | 28.2% | (33) |  |
| mRNA unspecified | 5.3% | (11) | 0.0% | (0) | 2.6% | (3) |  |
| Disease Isotype |  |  |  |  |  |  | 0.6880 |
| IgG | 57.2% | (119) | 58.3% | (13) | 54.7% | (64) |  |
| IgA | 23.1% | (48) | 33.3% | (4) | 26.5% | (31) |  |
| Light chain disease only | 17.8% | (37) | 8.3% | (5) | 15.4% | (18) |  |
| Other | 2.4% | (5) | 0.0% | (0) | 3.4% | (4) |  |
| SMM | 5.3% | (11) | 0.0% | (0) | 4.3% | (5) | 1.0000 |
| Active MM |  |  |  |  |  |  |  |
| Time since diagnosis (months) | 60 | [0-254] | 60 | [0-187] | 61 | [0-238] | 0.4904 |
| > 3 previous lines of treatment | 27.4% | (57) | 45.5% | (10) | 22.2% | (26) | **0.0292** |
| Disease response status: |  |  |  |  |  |  |  |
| CR or sCR | 43.3% | (90) | 22.7% | (5) | 49.6% | (58) | **0.0339** |
| VGPR | 22.1% | (46) | 31.8% | (7) | 20.5% | (24) | 0.2680 |
| PR or MR | 9.1% | (19) | 13.6% | (3) | 6.8% | (8) | 0.3805 |
| SD or PD | 14.4% | (30) | 22.7% | (5) | 12.0% | (14) | 0.1847 |
| Unable to assess | 11.1% | (23) | 9.1% | (2) | 11.1% | (13) | 1.0000 |
| Neutropenia ≥ G3 (< 1,500/μL) | 2.0% | (4/197) | 0.0% | (0) | 1.8% | (2/109) | 1.0000 |
| Lymphopenia ≥ G3 (< 800/μL) | 9.7% | (19/196) | 36.4% | (8) | 7.4% | (8/108) | **0.0011** |
| Immunoparesis | 88.2% | (181/205) | 95.5% | (21) | 87.9% | (102/116) | 0.4650 |
| Treatment regimen contains: |  |  |  |  |  |  |  |
| Immunomodulatory drug | 46.6% | (97) | 45.5% | (10) | 49.6% | (58) | 0.8178 |
| Proteasome inhibitor | 27.9% | (58) | 36.4% | (8) | 29.9% | (35) | 0.6170 |
| Steroid | 49.5% | (103) | 59.1% | (13) | 48.7% | (57) | 0.4867 |
| Anti-CD38 mAb | 45.7% | (95) | 59.1% | (13) | 45.3% | (53) | 0.2540 |
| BCMA-targeted therapy | 11.5% | (24) | 36.4% | (8) | 6.8% | (8) | **0.0006** |
| BCMA-targeted bispecific | 3.4% | (7) | 22.7% | (5) | 0.0% | (0) | **<0.001** |
| CAR-T cell therapy | 8.2% | (17) | 13.6% | (3) | 6.0% | (7) | 0.1957 |
| CAR-T <12 mo before dose 1 | 3.4% | (7) | 9.1% | (2) | 1.5% | (2) | 0.1179 |
| Other bispecific (non-BCMA) | 4.3% | (9) | 4.5% | (1) | 5.1% | (6) | 1.0000 |
| Other therapy (incl. venetoclax, selinexor, elotuzumab, alkylators) | 13.9% | (29) | 18.2% | (4) | 12.0% | (14) | 0.7336 |
| Previous ASCT | 53.8% | (112) | 50.0% | (11) | 53.8% | (63) | 0.8177 |
| No active treatment | 16.3% | (34) | 0.0% | (0) | 17.1% | (20) | **0.0430** |

**Note:** values are presented as percentage (n) or median [range]; p-values according to Mann-Whitney U test for continuous variables and Fisher's exact test for categorical variables. Lab values, disease response status and treatment regimen were registered at the date of administration of the first dose of mRNA vaccine.
**Abbreviations:** Ig, immunoglobulin; MM, multiple myeloma; SMM, smoldering multiple myeloma; CR, complete response; sCR, stringent complete response; VGPR, very good partial response; PR, partial response; MR, minimal response; SD, stable disease; PD; progressive disease; G3, grade 3 according CTCAE v5.0; G2, grade 2 according CTCAE v5.0; ANC, absolute neutrophil count; ALC, absolute lymphocyte count; ASCT, autologous stem cell transplant; mAb, monoclonal antibody; BCMA, B-cell maturation antigen; CAR, chimeric antigen receptor.

Of the 139 fully vaccinated patients for whom SARS-CoV-2 antibody titers were available >10 days after the second dose, 117 (84.2%) mounted a measurable SARS-CoV-2 IgG antibody response (i.e. > 5 AU/mL, median 208 AU/mL, range: 7-7882 AU/mL). The 118 MM patients without COVID-19 prior to vaccination had 3.6-fold lower antibody levels compared to a fully vaccinated age- and gender-matched control group of health care workers who were all seronegative at the time of vaccination (“control group”) (median of 77 AU/mL vs. 279 AU/mL, p=0.001, Figure 2A). Notably, antibody levels in the 21 MM vaccine responders that did have evidence of prior COVID-19 were 4.3-fold higher compared to the control group (median of 1199 AU/mL vs. 279 AU/mL, p<0.001, Figure 2A). Patients receiving MM treatment had significantly lower anti-spike IgG antibody levels after two doses (Supplementary Figure S7L, p=0.0084) compared to MM patients not on treatment. Looking at treatment categories, we found significantly lower antibody levels for patients receiving anti-CD38-containing regimens (p<0.001) and BCMA-targeted therapy (p=0.002) but not for all other treatments (p=0.95) compared to patients that are not actively being treated (Figure 2B). Other significant clinical factors that could influence response to vaccination are shown in Supplementary Figure S7.

Of note, 15.9% of MM patients (22/139) failed to develop SARS-CoV-2 IgG antibodies above the limit of test detection despite having received both doses of mRNA vaccines. We note that 20 of the 22 MM patients that did not develop antibodies were receiving treatment containing either a BCMA-targeted drug or an anti-CD38 monoclonal antibody. Time course of the antibody levels confirmed delayed and suboptimal responses in the majority of MM patients without prior SARS-CoV-2 infection (Figure 2C, Supplementary Figures S5-6).

Univariate analysis showed a significant association of the following factors with absence of anti-spike IgG after full mRNA vaccination: >3 previous lines of treatment (p=0.029), receiving active MM treatment (p=0.0430), absence of (stringent) complete response status (sCR/CR) (p=0.034), grade 3 lymphopenia (p=0.001) and receiving BCMA-targeted therapy (p<0.001) (Table 1). Multivariate logistic regression found that, after correcting for age, vaccine type, lines of treatment, time since MM diagnosis, response status and lymphopenia, anti-CD38-containing treatment was borderline non-significant (p=0.066, OR=5.098) but BCMA-targeted treatment remained significantly associated with the probability of not developing antibodies after vaccination (p=0.002, OR=32.043) (Supplementary Table S1).

## DISCUSSION

In our report, patient A died of COVID-19 after being vaccinated with both doses of mRNA vaccine without mounting serologic or cellular adaptive immunity to SARS-CoV-2. Given the recent FDA approvals of a BCMA-targeted antibody-drug conjugate^9,10^ and CAR-T therapy^11,12^, BCMA-targeted agents are increasingly going to be adopted into standard management of myeloma patients. There is, therefore, an urgent need to study timing of vaccination, SARS-CoV-2 infection prophylaxis and post-vaccine management in this patient population.

There may be a temporal relationship between BCMA-directed therapy and vaccine efficacy that warrants further research, exemplified by patient A who received the vaccine within 3.5 months of CAR-T therapy and did not mount measurable immune responses. In contrast, patient C who got vaccinated 18 months after CAR-T infusion, had a robust serological response with demonstrated B and T cell activity against SARS-CoV-2. For MM patients undergoing autologous stem cell transplant, we wait 12 months before initiating routine vaccinations to maximize efficacy^13,14^. We counsel MM patients at our institution to receive COVID-19 vaccination per CDC guidelines^15-17^. For the subgroup of patients receiving BCMA-targeted CAR-T treatment, we defer COVID-19 vaccination for three months after CAR-T based on published international guidelines^6^. This may have to be revisited as more data is made available for this patient population.

Our report provides data of the largest cohort of MM patients with serological response measurements to SARS-CoV-2 mRNA vaccines after completed mRNA SARS-CoV-2 vaccination to date. Our finding that over 15% of fully vaccinated MM patients do not have a detectable serologic response, combined with our experience of severe and fatal COVID-19 in one such non-responder highlight the critical need to monitor vaccine response in this high risk population. Non-responders may need to continue public health measures to protect themselves as the pandemic continues and may benefit from study of prophylactic antibody treatments to avoid contracting SARS-CoV-2 or to attenuate disease. We further found that patients with MM on multiple (>3) treatments, current BCMA-targeted treatment and lymphopenia were more likely to have absent antibody response and treatment regimens including anti-CD38 monoclonal antibody led to significantly lower levels of anti-spike IgG. The cohort may contain an overrepresentation of patients on BCMA-targeted treatments as we are a tertiary care center for MM where patients are routinely referred for treatment on clinical trials. Given the risk for developing COVID-19 and need for maintaining masking and other precautionary measures, our findings underscore the need for additional prospective serological monitoring in this subset of MM patients following COVID-19 vaccination.

In the patients that we studied extensively so far, absence of the CD19+ B cell compartment in the peripheral blood was associated with lack of antigen-specific B and also T cell responses. B cell depletion after anti-CD38 monoclonal antibody treatment regimens has been reported^18-20^, but the effects of BCMA-targeted treatment modalities on B cell populations (and the general immune composition) is less clear. B cell counts could be measured relatively easily and immunophenotyping in a larger cohort is warranted for independent confirmation of our findings. Furthermore, the mechanism by which BCMA-targeted treatment contributes to lack of T cell vaccine responses requires further in-depth studies since there is no obvious explanation. Reports have shown that, after SARS-CoV-2 infection, T cell responses can be present, even in the absence of circulating antibodies^7,21^. How this translates to the post-vaccine setting is unclear, especially in patients that are immunocompromised. If B cell counts are indeed transiently reduced after BCMA-targeted treatment, as our findings suggest, it might be beneficial to consider a bridging strategy to protect patients during the critical time before a serological response against SARS-CoV-2 can be mounted.

The relatively high percentage of vaccine non-responders in the MM population and the potential for cancer-directed therapies to hamper vaccine responses more broadly, support the need for serological monitoring of vaccine responses using quantitative assays, ongoing public health protective measures to avoid contracting SARS-CoV-2, and further study of prophylactic or early treatment modalities against COVID-19 in this high-risk population.

## Data Availability

All requests for raw and analyzed data and materials will be promptly reviewed by the Icahn School of Medicine at Mount Sinai and the Mount Sinai Hospital to verify if the request is subject to any confidentiality and data protection obligations. Any data and materials that can be shared will be released via a material transfer agreement.

## AUTHOR CONTRIBUTIONS

VS, AW, SP and the PVI study group provided conceptualization, methodology, analysis and resources for this work. SA, KK, KB, KS, CRG were involved in organizational aspects of the clinical studies, patient recruitment, data collection and analysis. OVO, SA, KS, CRG, BW, THM, EMS, FK and CCC were involved in design, data collection, analysis, visualization and interpretation of serological data. AA, BU, KT, DG, AR, SKS and SG were involved in design, execution, analysis, visualization and interpretation of T and B cell assays. ZK, ASGR and HvB were involved in execution, analysis and interpretation of genomic data. NWS, KM, AWC, AR, SKS, SG and MM were involved in design, data collection and analysis of the Mount Sinai COVID-19 biobank data. LS, SJ, AC and SJ were involved in different aspects of patient care. NB, SG, MM, DBD, SJ, VS, AW and SP provided interpretation of the data and conceptualization of the first manuscript draft. OVO, AA, BU, VS, AW and SP contributed to the writing of the first manuscript draft. All coauthors provided critical edits to the initial manuscript draft and approved the final version.

## ACKNOWLEDGEMENTS

We would like to thank the study participants for their generosity and willingness to participate in longitudinal COVID19 research studies. We would like to acknowledge the clinical and research staff at the Center of Excellence for Multiple Myeloma at Mount Sinai for their help. We acknowledge the lab of Alessandro Sette at the La Jolla Institute for Immunology and the lab of Steven C. Almo at Albert Einstein College of Medicine for sharing reagents that were used in this work.

PVI study group: Bulbul Ahmed, Hala Alshammary, Dr. Deena Altman, Angela Amoako, Mahmoud Awawda, Carolina Bermúdez-González, Rachel Chernet, Lily Eaker, Shelcie Fabre, Emily. D. Ferreri, Daniel Floda, Dr. Giulio Kleiner, Dr. Denise Jurczyszak, Julia Matthews, Wanni Mendez, Dr. Lubbertus CF Mulder, Jose Polanco, Kayla Russo, Ashley Salimbangon, Dr. Miti Saksena, A. Shin, Levy Sominsky, Sayahi Suthakaran

## SUPPLEMENTARY FIGURE LEGENDS

**Supplementary Figure S1:**
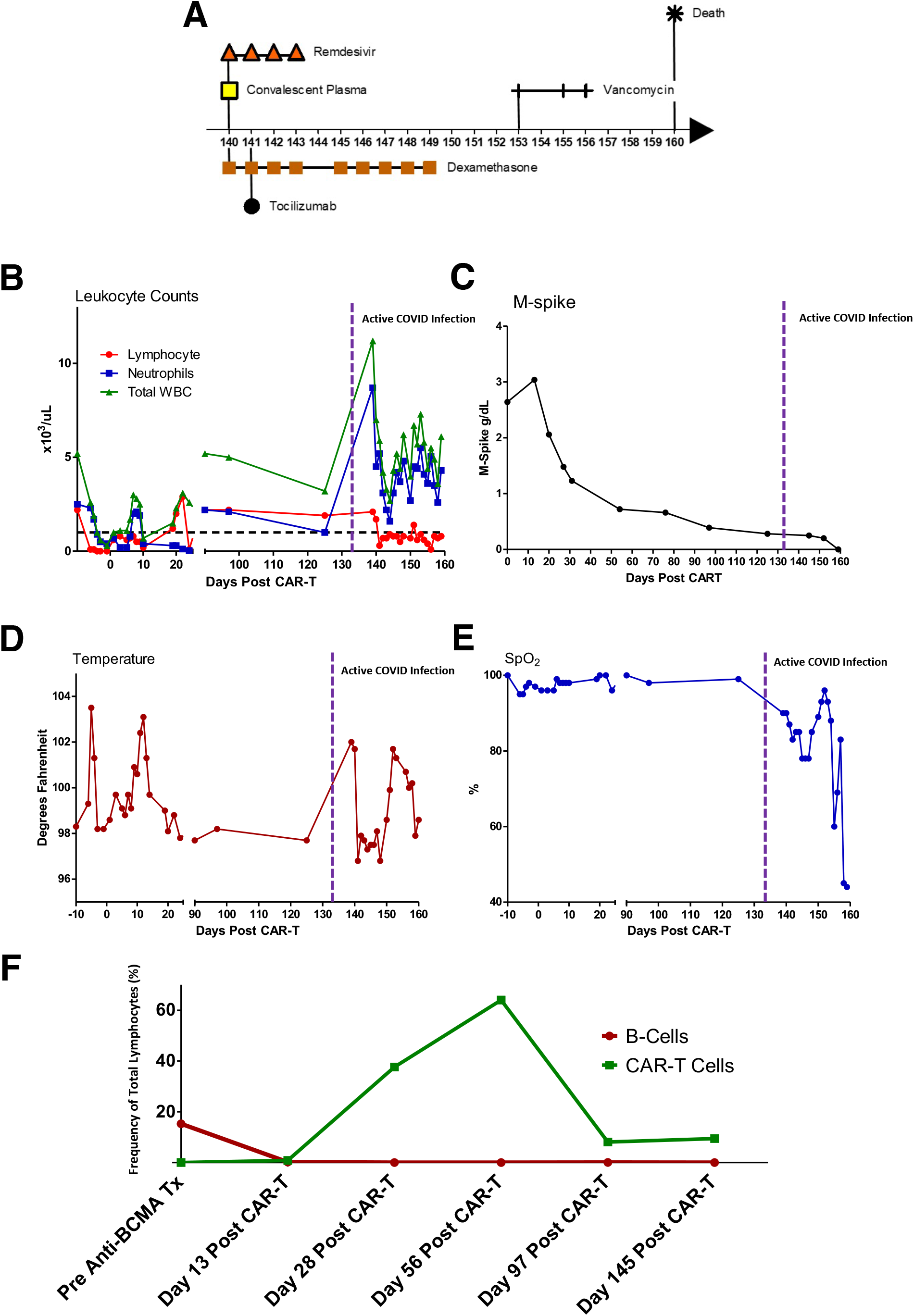
Detailed clinical course and parameters during the time period after chimeric antigen receptor (CAR)-T therapy. (A) Clinical event timeline showing relevant COVID-19-related interventions during ICU admission. (B) Leukocyte count evolution shown on a timeline starting at CAR-T infusion (day 0). Graph shows total white blood cell (WBC) count (green), absolute neutrophil count (blue) and absolute lymphocyte count (red). Vertical line illustrates start date of symptomatic COVID-19 infection. Horizontal line is reference line of normal absolute lymphocyte count. (C) M-spike evolution shown on a timeline starting at CAR-T infusion (day 0). Vertical line illustrates start date of symptomatic COVID-19 infection. (D) Temperature curve shown on a timeline starting at CAR-T infusion (day 0). Vertical line illustrates start date of symptomatic COVID-19 infection. (E) Peripheral oxygen saturation shown on a timeline starting at CAR-T infusion (day 0). Vertical line illustrates start date of symptomatic COVID-19 infection. (F) Timeline showing B cells (red) and CAR-T cells (green) as a fraction of total lymphocytes.

**Supplementary Figure S2:**
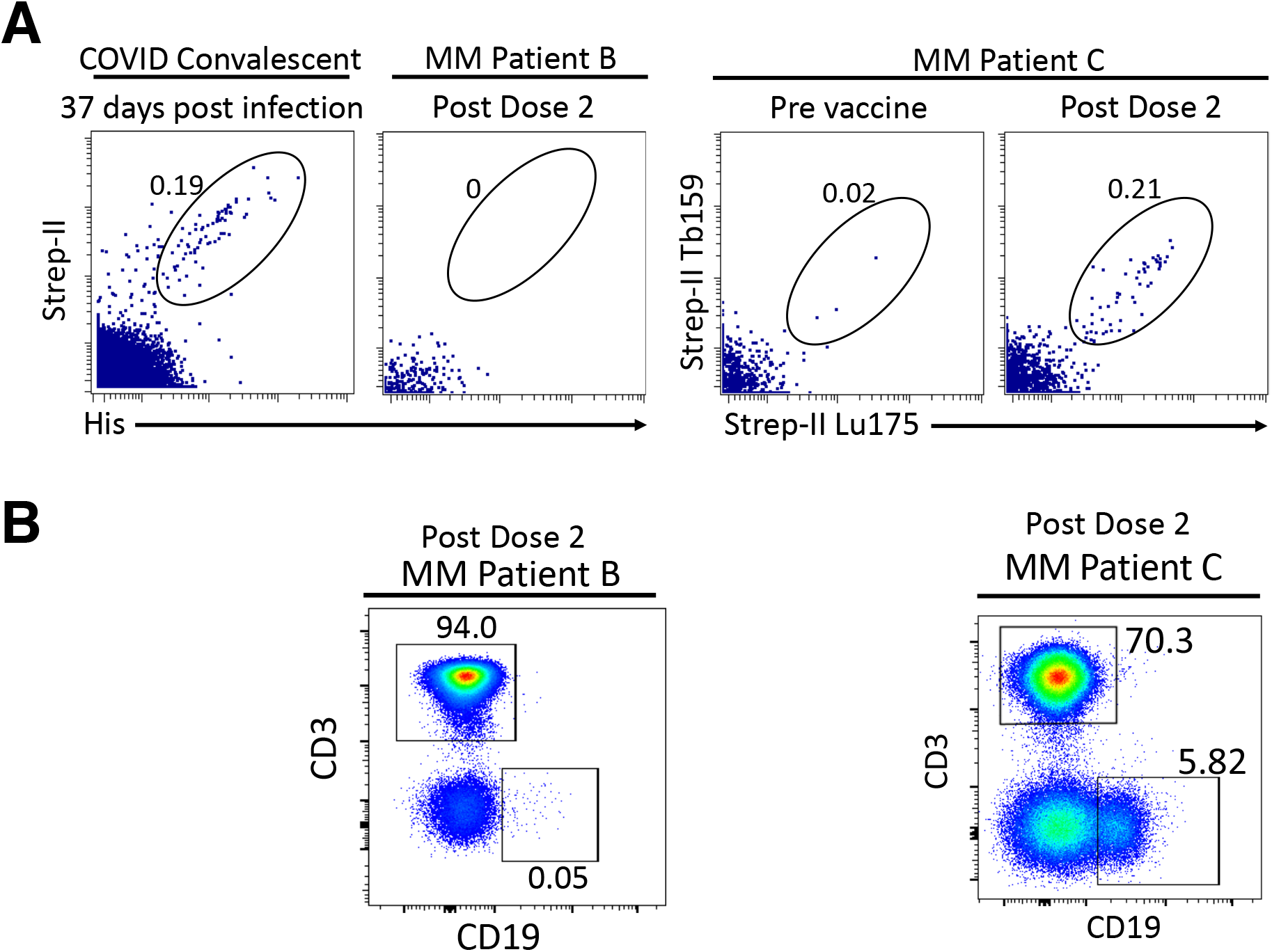
Presence of SARS-CoV-2 spike-specific B cells and T and B cell distribution in overall lymphocyte population. (A) Presence/absence of spike-positive (i.e. spike-specific) B cells by flow cytometry (% of CD19+ cells shown). Presence of spike-positive B cells shown in a COVID-19 convalescent patient 37 days after the start of molecularly confirmed COVID-19 infection. Absence of spike-positive B cells for multiple myeloma (MM) patient B (58 days after vaccination) shown on the second plot from the left side. Absence of spike-positive B cells in MM patient C (43 days before vaccine dose 1) and appearance of spike-positive B cells in the same patient (6 days after vaccine dose 2) shown on the right side. Numbers on dot plot represent percentage of spike-positive B cells within the total B cell gate. (B) Severe depletion of B-cells in patient undergoing treatment with a regimen containing anti-CD38 monoclonal antibody post vaccine dose 2, in contrast to patient C who has B cells present one year after CAR-T infusion (sample taken 6 days after vaccine dose 2). Numbers on dot plot represent frequencies of total T (CD3+) and B (CD19+) cells within the lymphocyte gate.

**Supplementary Figure S3:**
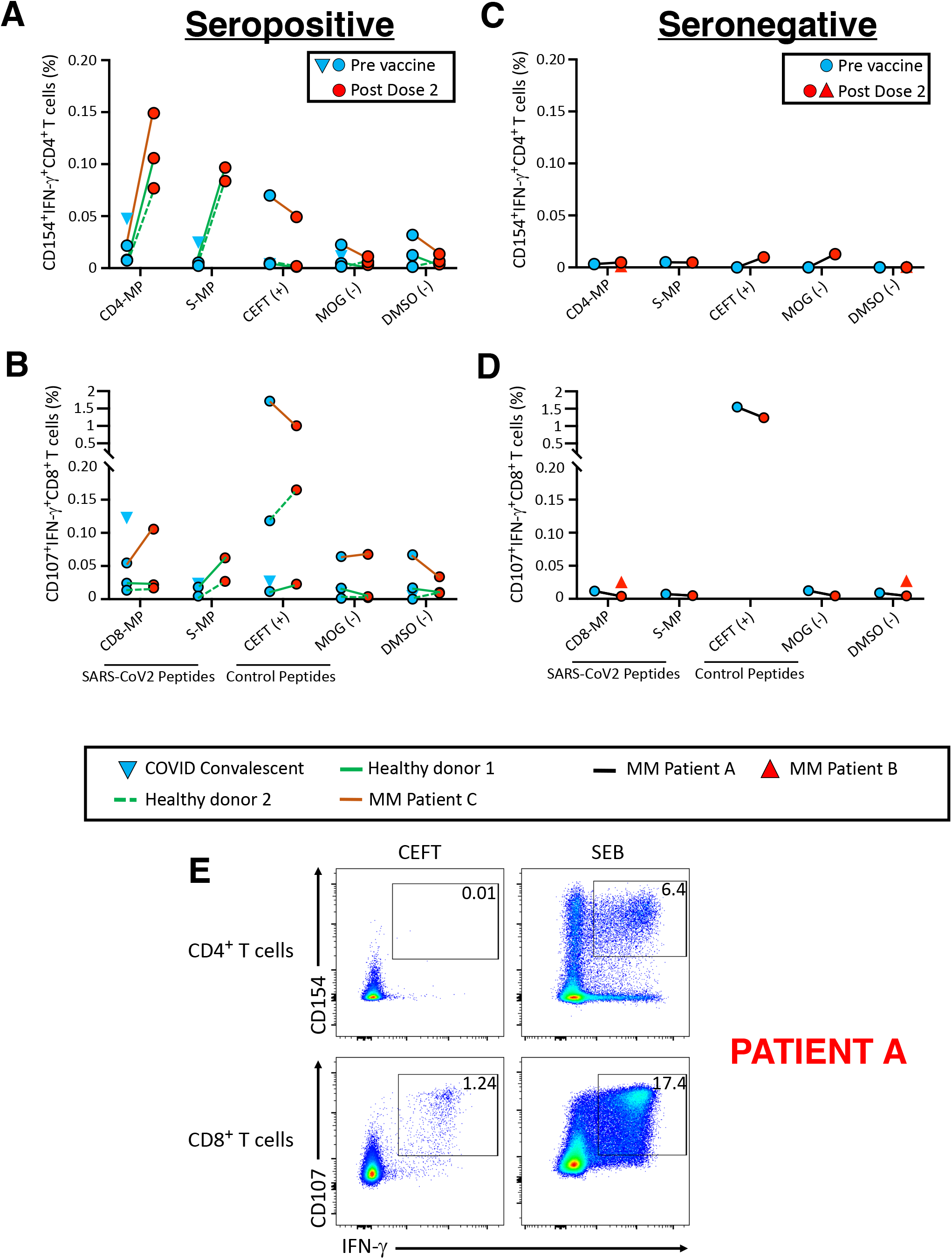
SARS-CoV-2 specific T cell responses in multiple myeloma (MM) patients, vaccinated healthy controls and a COVID-19 convalescent healthy donor measured by intracellular cytokine flow cytometry assay. Peripheral blood mononuclear cells (PBMC) were cultured with either SARS-CoV-2 peptides pools (CD4-MP, CD8-MP-S-MP), positive control CEFT peptide pool, negative control MOG peptide pools or DMSO. Frequencies of IFN-γ-secreting activated CD4^+^ T cells (CD4^+^CD154^+^IFN-γ^+^) or IFN-γ-secreting activated CD8+ T cells (CD8^+^CD107^+^IFN-γ^+^) under different stimulation conditions measured by Flow Cytometry (% of CD3^+^CD4^+^ T cells or % of CD3^+^CD8^+^ T cells shown). (A,C) Change in the frequency of IFN-γ-secreting activated CD4+ T cells prior to receiving mRNA vaccine compared to a time point after vaccine dose 2 in the seropositive (i.e. with detectable titers of anti-spike IgG after vaccination or infection) (A) and seronegative (i.e. without detectable titers of anti-spike IgG) (C) group (% of CD3+CD4+ T cells shown. (B,D) Change in the frequency of IFN-γ-secreting activated CD8+ T cells (CD8+CD107+IFN-γ+) prior to receiving mRNA vaccine compared to a time point after vaccine dose 2 in the seropositive (B) and seronegative (D) group (% of CD3+CD8+ T cells shown). Lines denote different patients while symbols illustrate time point in relation to vaccine dose. (E) Flow cytometry dot plots of T cells of patient A, demonstrating functional T cell responses as measured by the presences of IFN-γ-secreting activated CD4^+^ and CD8^+^T cells in response to activation by CEFT and SEB. Numbers denote frequencies of IFN-γ-secreting cells in total CD4^+^ and CD8^+^ T-cell population.

**Supplementary Figure S4:**
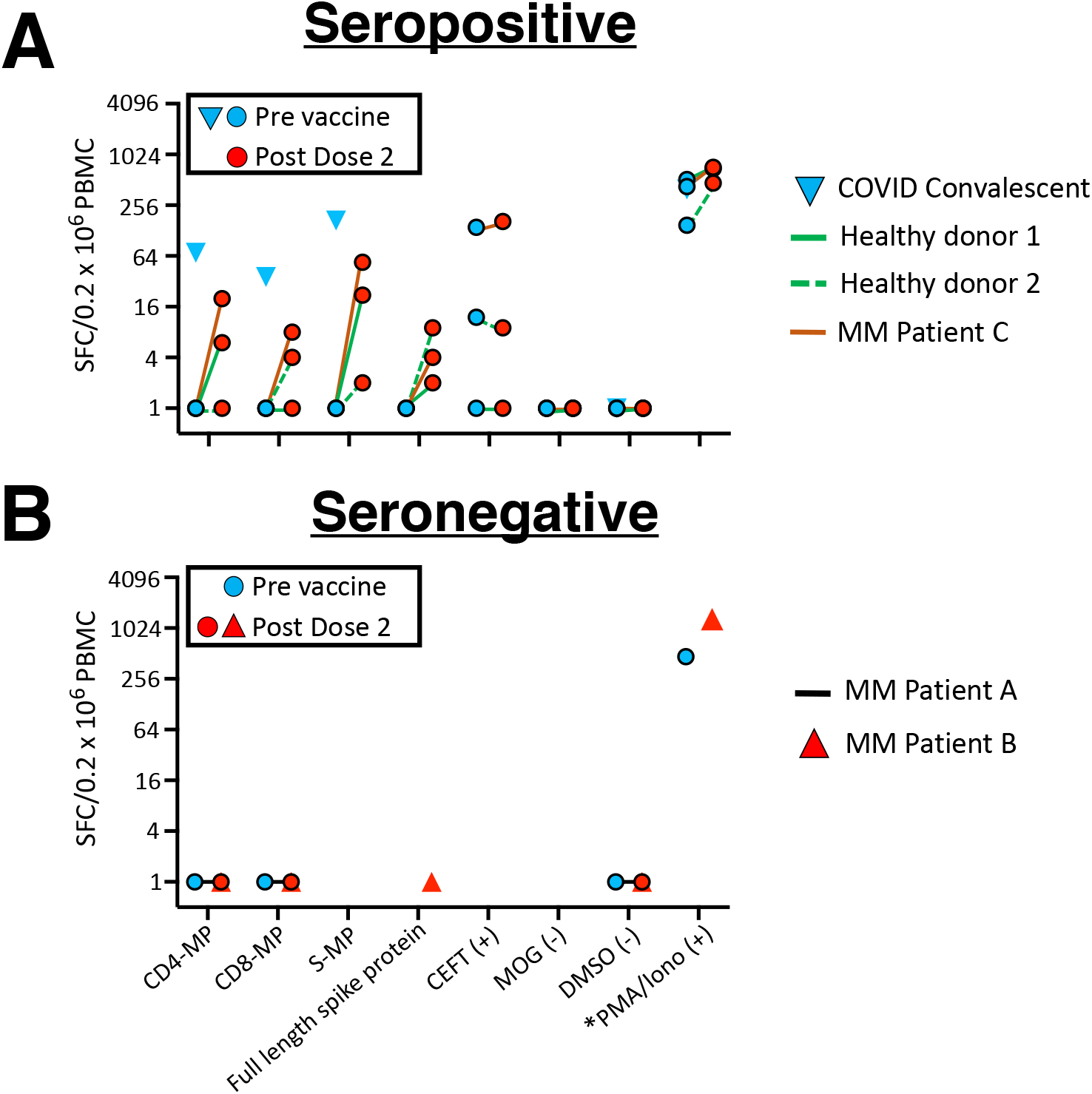
SARS-CoV-2 specific T cell responses in multiple myeloma (MM) patients, vaccinated healthy donors and a COVID-19 convalescent healthy donor measured by Elispot assay. Peripheral blood mononuclear cells (PBMC) were plated at 200,000 (200K) with either SARS-CoV-2 peptides pools (CD4-MP, CD8-MP-S-MP), positive control CEFT peptide pool, negative control MOG peptide pools or DMSO. IFN-γ+ spot forming cells (SFC) were quantified for each condition. (A) Change in IFN-γ+ spots from pre-vaccine to after second dose of vaccine in seropositive subjects (i.e. with detectable titers of anti-spike IgG after vaccination or infection). (B) Lack of in IFN-γ+ spots from seronegative (i.e. without detectable titers of anti-spike IgG) patient A and patient B. Positive control PMA/IONO was plated at 20K to allow on-scale comparison. For visualization purposes 0 spots are plotted as 1 SFC.

**Supplementary Figure S5:**
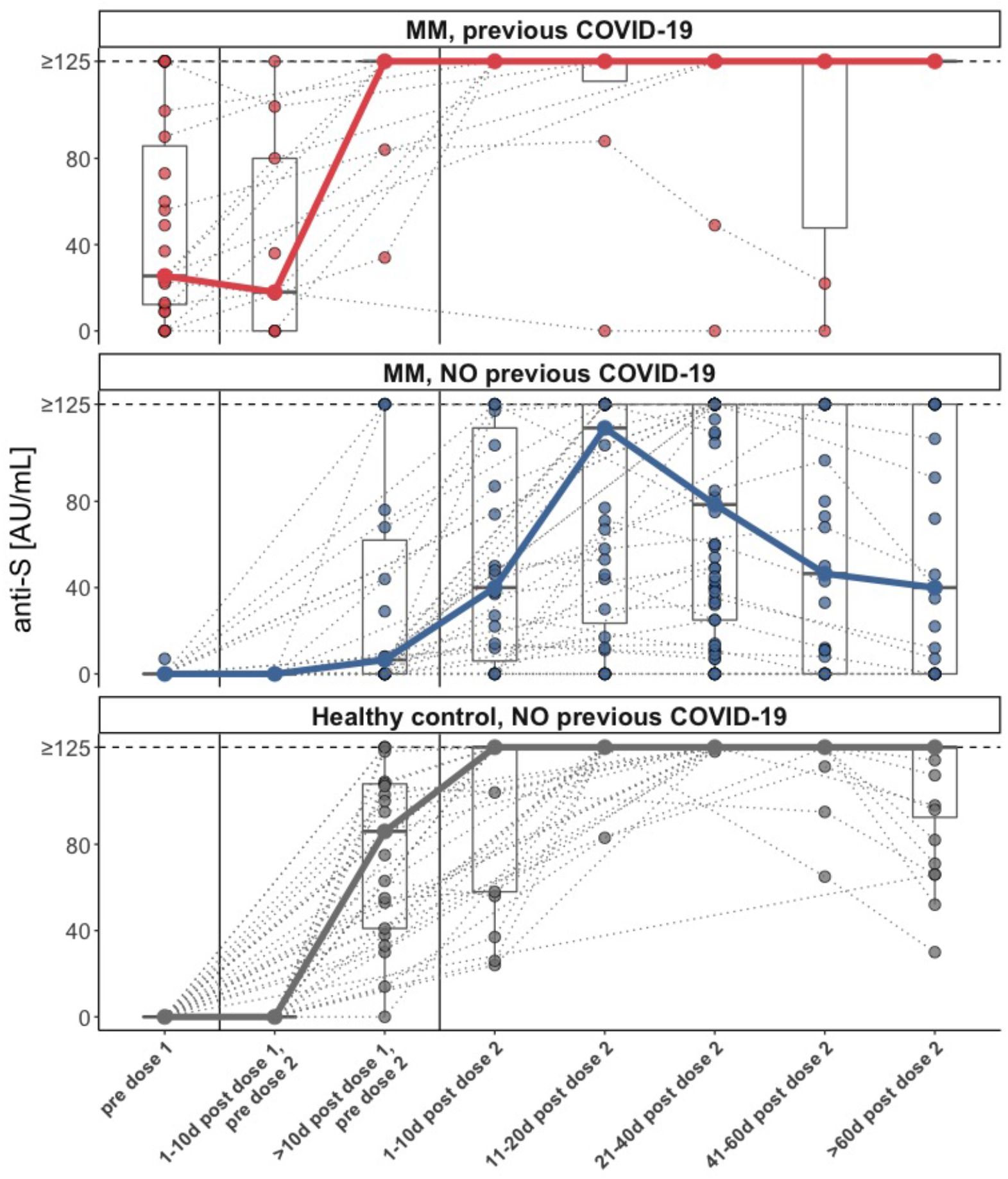
Time course of SARS-CoV-2 anti-spike (S) IgG antibody levels in healthy donors versus myeloma patients with/without previous COVID-19 infection. Time course of SARS-CoV-2 anti-S IgG antibody levels (shown capped at 125 AU/mL) in multiple myeloma (MM) patients with prior COVID-19 infection (red, top), in MM patients without prior COVID-19 infection (blue, center) and in healthy controls without prior COVID-19 infection (gray, bottom). Measurements from the same study participant are linked with dotted lines. Thick lines connect the median at each time point.

**Supplementary Figure S6:**
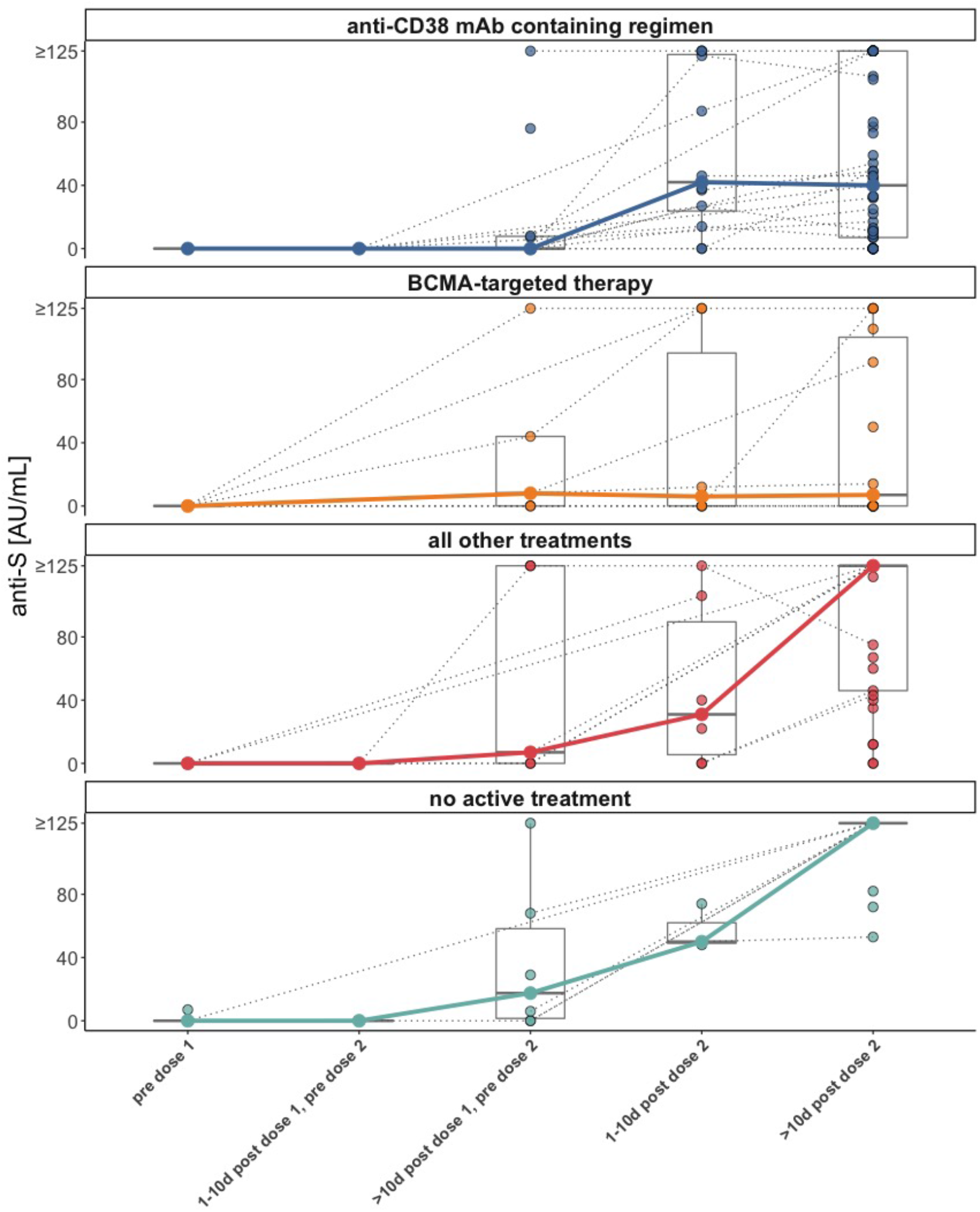
Time course of SARS-CoV-2 anti-spike (S) IgG antibody levels in myeloma patients split according to major treatment groups. Time course of SARS-CoV-2 anti-S IgG antibody levels (shown capped at 125 AU/mL) in multiple myeloma (MM) patients treated with a anti-CD38 monoclonal antibody (mAb)-containing regimen (blue, top), MM patients treated with a BCMA-targeted therapy (second row, orange), MM patients treated with all other treatments (third row, red) and MM patients that were not receiving active treatment at the time of vaccination (bottom, teal).

**Supplementary Figure S7:**
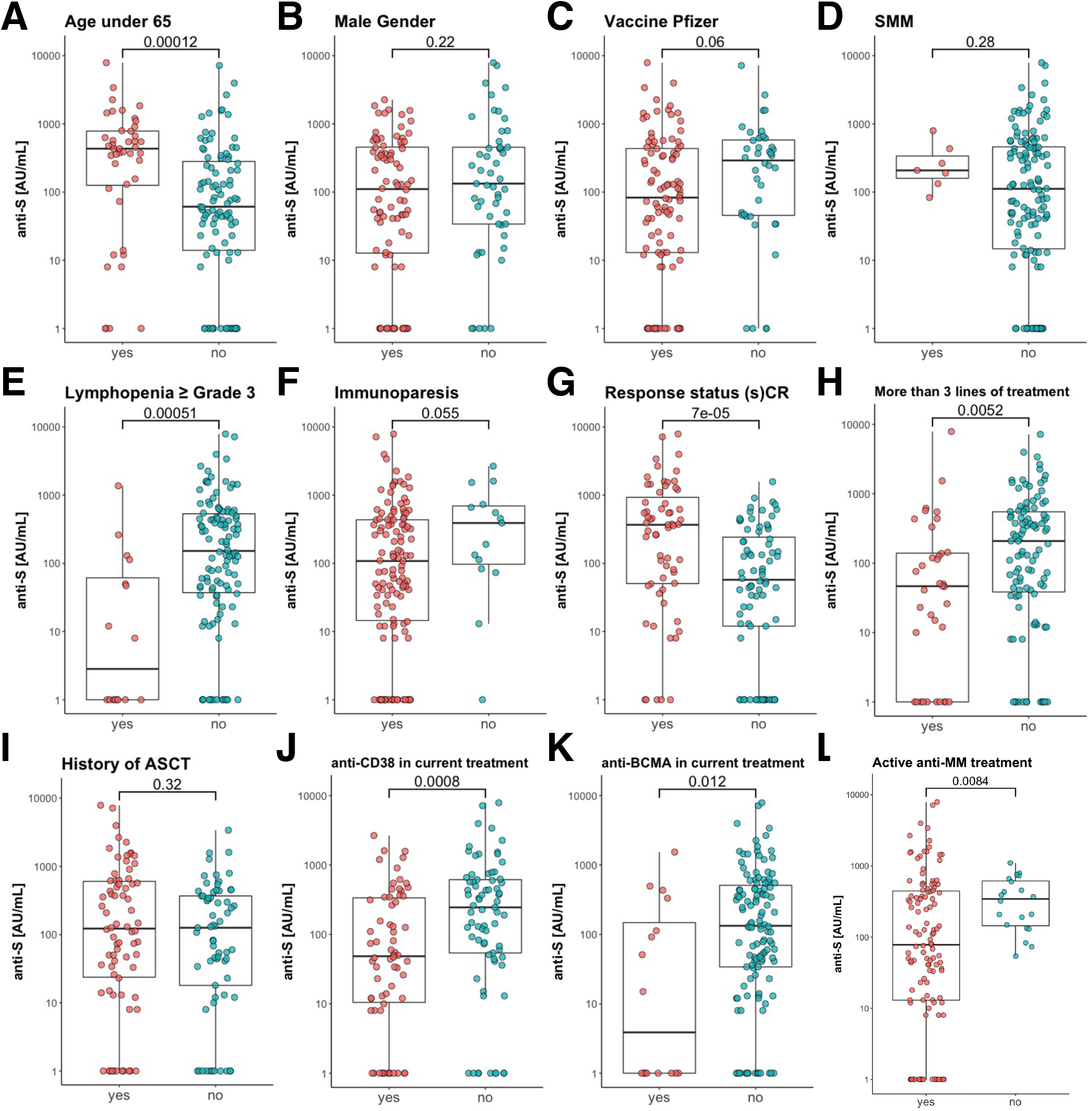
Factors univariately associated with levels of SARS-CoV-2 anti-spike (S) IgG antibody levels in patients with multiple myeloma (MM) more than ten days after receiving two doses of mRNA vaccine. Boxplots with overlaying jitter plots illustrating the association of different clinical and treatment characteristics with the level of anti-S IgG more than ten days after receiving two doses of mRNA SARS-CoV-2 vaccine. Shown are age less than 65 years (A); male gender (B); vaccine type Pfizer-BioNTech (C); smoldering multiple myeloma (SMM) diagnosis (D); lymphopenia ≥ grade 3 (i.e. absolute lymphocyte count < 500/µL) (E); immunoparesis (i.e. levels of one or more uninvolved immunoglobulin subtypes below normal) (F); response status according to International Myeloma Working Group (IMWG) criteria of stringent complete response (sCR) or complete response (CR) (G); having received more than 3 previous lines of treatment (H); history of autologous stem cell transplant (ASCT) (I); anti-CD38 monoclonal antibody as a part of the current treatment (J); BCMA-targeted treatment as a part of the current treatment (K); and receiving active anti-MM treatment at the time of vaccination (L). P-values represent comparison using the non-parametric Mann-Whitney U test.

**SUPPLEMENTARY TABLE S1.**
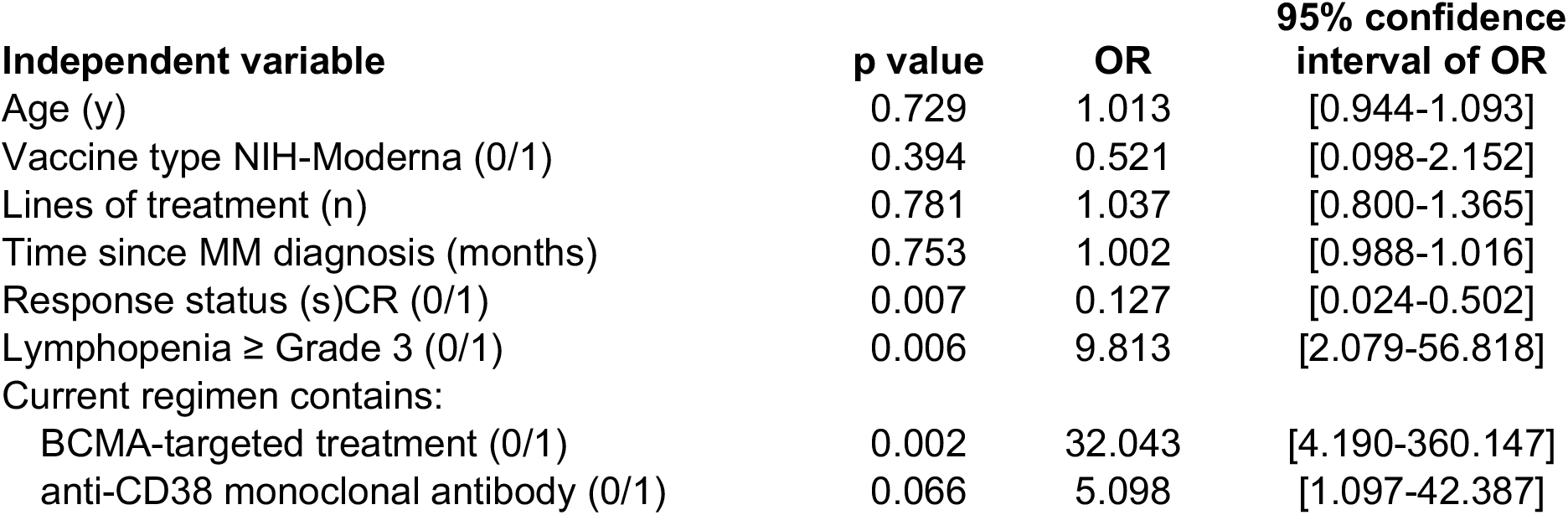
Multivariate logistic regression model with absence of detectable IgG antibody levels > 10 days after full vaccination as dichotomized outcome.

