## Supplementary Data for "Fatal breakthrough infection after anti-BCMA CAR-T therapy highlights suboptimal immune response to SARS-CoV-2 vaccination in myeloma patients"

### MATERIALS AND METHODS

**PATIENT SELECTION:** The study cohort consisted of 208 patients with and without previously documented COVID-19 pooled from four different Institutional Review Board (IRB) approved protocols at The Icahn School of Medicine at Sinai.

A total of 77 MM patients were enrolled after obtaining written informed consent for an ongoing longitudinal study at our institution (IRB-16-00791). Patients had blood and saliva taken for analysis at multiple time points around the administration of the SARS-CoV-2 mRNA vaccine starting at pre-vaccine to post vaccine. All specimens were coded prior to processing and antibody testing for all serum specimen was performed in a blinded manner. All participants with, at least, one post vaccine antibody data point available at the time of writing this report were included in the analysis.

The remaining 131 MM patients were identified under a retrospective study (IRB: GCO#: 11-1433) by conducting a chart review for patients at our MM clinic who had SARS-CoV-2 spike IgG results at various time points around SARS-CoV-2 mRNA vaccine administration. Both studies were carried out in compliance with the Declaration of Helsinki and International Conference on Harmonization Guidelines for Good Clinical Practice. Chart review was conducted to retrieve patient clinical characteristics. Anti-SARS-CoV-2 antibody testing was performed using an anti-IgG assay developed at Mount Sinai Health System Department of Pathology in collaboration with the Icahn

School of Medicine at Mount Sinai Department of Microbiology under a Food and Drug Administration (FDA) Emergency Use Authorization.

Control group: 38 participants of the PARIS (***P****rotection* ***A****ssociated with* ***R****apid* ***I****mmunity to* ***S****ARS-CoV-2*) study were selected as controls to best match the demographics of the MM patient population. This study follows health care workers over time to investigate the durability and effectiveness of SARS-CoV-2 immune responses in health care workers over time. The study was reviewed and approved by the Mount Sinai Hospital Institutional Review Board (IRB-20-03374). All participants provided informed consent prior to collection of data and specimen. All specimens were coded prior to processing and antibody testing for all serum specimen was performed in a blinded manner.

**SARS-CoV-2 antibody ELISA**: Antibodies to SARS-CoV-2 spike were detected using an established quantitative two-step ELISA termed Mount Sinai Antibody test described in detail in the referenced manuscripts[1,2]. The assay shows a performance of 100% specificity and 95% sensitivity in in-house evaluation.

**STATISTICAL ANALYSIS:** Continuous variables are presented throughout the manuscript as median and range. Categorical variables are shown as a percentage and the absolute number of subjects. Wherever two outcome groups are compared, Fisher's exact test was used to determine significance and where applicable odds ratios (ORs) were reported. The Mann-Whitney U test was used to determine significance for continuous variables. A two-sided alpha $< 0.05$ was considered statistically significant. All statistical analyses were done using R (v4.0.2). Multivariate statistical analysis was done using the presence or absence of measurable anti-spike IgG response > 10 days after dose 2 as a binary outcome variable and conducted using the glm(…, family = “binomial”) function in R (v4.0.2).

**SAMPLE COLLECTION FOR FLOW AND MASS CYTOMETRY:** The subjects consented to enrollment in HSM: 18-00456 Multiple Myeloma Biorepository and MARS clinical trial IRB: 16-00791.The study was approved by the Program for Protection of Human Subject an Institutional Review Board approved research study. Peripheral blood was collected in heparin anticoagulated green tops, BD Vacutainer CPT (Cat#362761), and (Cat#367985) (10 mL) via venipuncture according to trial schedule. Peripheral blood mononuclear cells (PBMC) were Ficoll density separated and cryopreserved by the Parekh Lab and the Human Immune Monitoring Center at the Icahn School of Medicine (HIMC). Cryopreserved PBMC samples were used to Flow Cytometry and Mass Cytometry analysis

**FLOW CYTOMETRY:** All T cell assays were carried out in X-VIVO 15™ (Lonza) supplemented with 10% human AB serum (R&D Systems). PBMCs were cultured in 96 well U bottom well plates at 1 x $10^6$ cells per well for 6 hours in the presence of either SARS-COV2 peptide or control peptide pools. All culture conditions also included co-stimulators anti-CD28 (clone) and anti-CD49d (clone) (Biolegend), activation markers CD154-PE (Clone 24-31, Biolegend) and CD107a-FITC (Clone H4A3, Biolegend) and Monensin (Biolegend). The CD4 and CD8 SARS-COV2 peptide mega pools (MP) used in T cell assays were shared to us by Dr. Alessandro Sette (La Jolla Institute for Immunology, La Jolla, CA, MTA Agreement AGR-23018) and have been described in their publications[3,4]. CD4-MP is a pool comprises of two large MPs, one of which is a pool of predicted HLA Class II epitopes spanning the entire SARS-COV2 protein minus spike protein (n=221 peptides) and the other which is a pool of 15mers overlapping by 10 amino acids (aa) (n=246 peptides) derived only from the spike glycoprotein. The CD8-MP is a pool of 2 large MP (total 628 peptides) of peptides derived from predicted HLA class I epitopes from the total SARS-COV2 protein. Each individual peptide pool was used a conc of 1μg/ml in the cultures. In addition to Sette's groups peptide pools we used another spike peptide pool (S-MP) shared to us by Dr. Nina Bharadwaj (Icahn School of Medicine at Mount Sinai, New York, NY). This peptide pool consisted of two pools of peptides (n= 249 peptides) from N and C terminal half of SARS-COV2 spike glycoprotein and were 15mers overlapping by 10aa. Each peptide in this spike MP was used at a final concentration of 0.5μM in the cultures. All SARS-COV2 peptides used in our study were from the Ancestral Wuhan strain of SARS-COV2. Positive control CEFT peptides MPs were synthesized at GenScript and used at a final concentration of 0.7μM. CEFT peptides are a pool of 27 peptides composed of defined HLA class I and HLA class II epitopes from Cytomegalovirus, Epstein-Barr virus, Influenza virus and Clostridium tetani. As an additional positive control, we activated the cells with SEB [5μg/ml] (Toxin Technologies). As negative controls we used equimolar amount of DMSO and peptides derived from human Myelin-oligodendrocyte glycoprotein (MOG)[used at 1μM final concentration]. The MOG peptides were 15mers overlapping by 11aa and were synthesized at GenScript. Post stimulation cells were washed and stained with Live/Dead Fixable Blue Dead Cell

Stain Kit (L23105, Thermofisher Scientific) and fixed with 4% formaldehyde. Fixed cells were permeabilized with BD per buffer (BD Biosciences) and stained with a cocktail of antibodies listed in Supplementary Table S2. The cells were acquired on BD LSR Fortessa (BD Biosciences) and the data was exported to Flowjo (BD Biosciences) and Cytobank[5] for final analysis.

**ELISPOT ASSAYS:** Enzyme-linked immunospot (ELISPOT) flat-bottomed, 96-well nitrocellulose plates (MAHA S4510; Millipore) were coated with IFN-γ mAb (2 μg/ml, 1-D1K; MABTECH, Stockholm) and incubated for 2 hours at 37°C. After washing with PBS, plates were blocked with 10% human AB serum for 2 hours at 37°C. Cells were washed, concentrated, plated into each well titrating down in two-fold dilutions starting from 200,000 PBMCs, in the presence of peptides, protein, and controls (Dimethyl sulfoxide (DMSO) and CEFT peptide pools were used as negative and positive controls, respectively, along with phorbol 12-myristate 13-acetate (PMA)/ionomycin) for 40 hours in X-VIVO-15 serum-free medium. After incubation, the plates were semi-automatically washed thoroughly with PBS, and IFN-γ mAb (0.2 μg/ml, 7-B6-1-biotin; MABTECH) was added to each well. After incubation for 2 hours at 37°C, plates were washed and developed with streptavidin-alkaline phosphatase (1 μg/ml; Roche) for 1 hour at room temperature. After washing, substrate (5-bromo-4-chloro-3-indolyl phosphate/NBT; Sigma-Aldrich) was added and incubated for 12-15 min. After washing, the dark-violet spots were evaluated using the C.T.L. Immunospot analyzer and software (Cellular Technology Limited). Additional information can be found in Somaiah N et al.[6]

**DETECTION OF SPIKE-SPECIFIC B CELLS BY MASS CYTOMETRY (CYTOF):** Full length SARS-CoV-2 spike protein (Wuhan Strain) was used to detect spike-positive B cells. Spike protein was produced in the laboratory of Dr. Steven C. Almo at the Albert Eisenstein College of Medicine and shared with us. The spike protein production method and characterization has been published[7]. Since this overexpressed spike protein had Strep II and His tags at the C terminal end, we used either a anti-Strep II-anti-Strep II (each conjugated to different metals) or a anti-Strep II-anti-His combination strategy to detect Spike bound to B cells. Thawed cells were labeled with Rh103 intercalator (Fluidgm) as a viability dye for 20 minutes, washed and stained on ice with anti-IgG along with Spike protein for 30 minutes. IgG- and spike-stained cells were washed further stained with the remaining CyTOF antibody cocktail listed in CyTOF panel 1 (Supplementary Table S3) along with either anti-Strep II-anti-Strep II or a anti-Strep II-anti-His combination of antibodies. Post staining cells were washed and fixed in 2.4% formaldehyde containing 125nM intercalator-Ir (Fluidgm) and 300nM OsO4 (ACROS Organics) along with Palladium barcoding reagents (CyTOF Cell-ID 20-Plex Palladium Barcoding Kit, Fluidigm) to enable pooling of different samples and timepoints for simultaneous acquisition. Stained cells for CyTOF acquisition were further washed with CAS buffer (Fluidigm), re-suspended in CAS buffer containing EQ normalization beads (Fluidigm) and acquired on CyTOF2 (Fluidigm). After acquisition the data was normalized using bead-based normalization algorithm in the CyTOF software (Fluidigm). Normalized data was debarcoded using methods and software developed in Gary Nolan's group at the Stanford University School of Medicine[8]. Normalized and debarcoded data was uploaded to Cytobank[5] for final analysis.

**B CELL SCREENING & CAR-T CELL SCREENING:** Thawed PBMC were initially stained with Live/Dead Fixable Blue Dead Cell Stain Kit washed and stained with surface phenotypic markers listed in Supplementary Table S4. Presence of BCMA specific CAR-T cells were detected by adding BCMA protein conjugated to FITC (BCA-HF254, AcrosBiosystems) to the antibody cocktail. The cells were acquired on a BD LSR Fortessa. Data was analyzed using Cytobank[5] & Flowjo.

### SUPPLEMENTARY TABLES

**SUPPLEMENTARY TABLE S1 – Multivariate logistic regression model with absence of detectable IgG antibody levels > 10 days after full vaccination as dichotomized outcome.**

| Independent variable | p value | OR | 95% confidence interval |
|---|---|---|---|
| Age (y) | 0.729 | 1.013 | [0.944-1.093] |
| Vaccine type NIH-Moderna (0/1) | 0.394 | 0.521 | [0.098-2.152] |
| Lines of treatment (n) | 0.781 | 1.037 | [0.800-1.365] |
| Time since MM diagnosis (months) | 0.753 | 1.002 | [0.988-1.016] |
| Response status (s)CR (0/1) | 0.007 | 0.127 | [0.024-0.502] |
| Lymphopenia ≥ Grade 3 (0/1) | 0.006 | 9.813 | [2.079-56.818] |
| Current regimen contains: | | | |
| BCMA-targeted treatment (0/1) | 0.002 | 32.043 | [4.190-360.147] |
| anti-CD38 monoclonal antibody (0/1) | 0.066 | 5.098 | [1.097-42.387] |

**SUPPLEMENTARY TABLE S2 – Antibodies for flow cytometry based T cell assay**

| Catalog # | Target | Fluorophore | Clone | Company |
|---|---|---|---|---|
| 506538 | IFNg | BV421 | B27 | Biolegend |
| 561640 | CD45RA | V500 | HI100 | BD Biosciences |
| 302236 | CD19 | BV570 | HIB19 | Biolegend |
| 500332 | IL-2 | BV605 | MQ1-17H12 | Biolegend |
| 310934 | CD69 | BV650 | FN50 | Biolegend |
| 302833 | CD27 | BV711 | O323 | Biolegend |
| 300472 | CD3 | BV785 | UCHT1 | Biolegend |
| 502312 | GM_GSF | PercpCy5.5 | BVD2-21C11 | Biolegend |
| 502930 | TNFa | PE-CY7 | MAB11 | Biolegend |
| 344614 | CD4 | APC | SK3 | Biolegend |
| 301822 | CD14 | Alexa700 | M5E2 | Biolegend |
| 301016 | CD8 | APC-CY7 | RPA-T8 | Biolegend |

**SUPPLEMENTARY TABLE S3 – Mass cytometry (CyTOF) antibody panel used for the detection of spike-specific B cells**

| CyTOF Panel | Vendor | Catalog Number | Clone |
|---|---|---|---|
| **Anti-human IgA – Cd112** | Southern Biotech | 2050-01 | Polyclonal |
| **Anti-human IgM – Cd114** | Biolegend | 314502 | MHM-88 |
| **Anti-human IgG – Cd116** | BD Biosciences | 555784 | G18-145 |
| **Anti-human CD45 – Y89** | Fluidigm | 3089003B | HI30 |
| **Anti-human CD45RB – Ln113** | Biolegend | 310202 | MEM-55 |
| **Anti-human CD11c – Ln115** | Biolegend | 337202 | BU15 |
| **Anti-human IgD – Pr141** | Biolegend | 348202 | IA6-02 |
| **Anti-human CD19 – Nd142** | Miltenyi | 130-122-301 | REA675 |
| **Anti-human CD145RA – Nd143** | Miltenyi | 130122-292 | REA562 |
| **Anti-human CD79b– Nd144** | BD Biosciences | 557592 | 3A2-2E7 |
| **Anti-human CD4 – Nd145** | Miltenyi | 130-122-283 | REA623 |
| **Anti-human CD8 – Nd146** | Miltenyi | 130-122-281 | REA734 |
| **Anti-human CD20 – Sm147** | Fluidigm | 3147001B | 2H7 |
| **Anti-human CD16 – Nd148** | Miltenyi | 130-108-027 | REA423 |
| **Anti-human CD127 – Sm149** | Fluidigm | 314011B | A019D5 |
| **Anti-human CD1c – Nd150** | Miltenyi | 130-122-298 | REA694 |
| **Anti-human CD123 – Eu151** | Miltenyi | 130-122-297 | REA918 |
| **Anti-human CD66b – Sm152** | Miltenyi | 130-108-019 | REA306 |
| **Anti-human CD25 – Eu153** | Biolegend | 356102 | M-A251 |
| **Anti-human CD86 – Sm154** | Biolegend | 305410 | IT2.2 |
| **Anti-human CD27 – Gd155** | Miltenyi | 130-122-295 | REA499 |
| **Anti-human PD-1 – Gd156** | Biolegend | 329926 | EH12.2H7 |
| **Anti-human CD5 – Gd158** | Biolegend | 300602 | UCHT2 |
| **Anti-Strep II – Tb159** | GenScript | A01732 | 5A9F9 |
| **Anti-human CD14 – Gd160** | Biolegend | 301802 | M5E2 |
| **Anti-human CD56 – Dy161** | Miltenyi | 130-108-016 | REA196 |
| **Anti-human CD23 – Dy162** | Biolegend | 338502 | EBVCS-5 |
| **Anti-human CXCR5 – Dy163** | Miltenyi | 130-122-325 | REA103 |
| **Anti-human CD40 – Dy164** | Biolegend | 334302 | HB14 |
| **Anti-human CD73 – Ho165** | Biolegend | 344002 | AD2 |
| **Anti-human CD24 – Er166** | Fluidigm | 3166007B | ML5 |
| **Anti-human CCR7 – Er167** | Fluidigm | 3167009A | G043H7 |
| **Anti-human CD3 – Er168** | Miltenyi | 130-122-282 | REA613 |
| **Anti-human CD71– Tm169** | Biolegend | 334102 | CY1G4 |
| **Anti-human CD38 – Er170** | Miltenyi | 130-122-288 | REA671 |
| **Anti-human CD95 – Yb171** | Biolegend | 305602 | DX2 |
| **Anti-human CD39 – Yb172** | Biolegend | 328202 | A1 |

| | | | |
|---|---|---|---|
| **Anti-human CXCR3 – Yb173** | Miltenyi | 130-108-022 | REA232 |
| **Anti-human HLADR – Yb174** | Miltenyi | 130-122-299 | REA805 |
| **Anti-Strep II – Lu175** | GenScript | A01732 | 5A9F9 |
| **Anti-His – Lu175** | Biolegend | 362615 | J095G46 |
| **Anti-human CD21 – Yb176** | Biolegend | 354902 | BU32 |
| **Anti-human CD11b – Bi209** | Fluidigm | 3209003B | ICRF44 |

**SUPPLEMENTARY TABLE S4 – Antibodies used for screening B and CAR-T cells**

| Catalog # | Target | Fluorophore | Clone | Company |
|---|---|---|---|---|
| 317434 | CD4 | BV421 | OKT4 | Biolegend |
| 561121 | CD19 | V500 | HIB19 | BD Biosciences |
| 344842 | CD3 | BV785 | SK7 | Biolegend |
| 344742 | CD8 | BV605 | SK1 | Biolegend |
